# Severe Acute Respiratory Syndrome Coronavirus 2 (SARS-CoV-2) Household Transmission during the Omicron Era in Massachusetts: A Prospective, Case-Ascertained Study using Genomic Epidemiology

**DOI:** 10.1101/2024.02.05.24302348

**Authors:** Jaspreet Banga, Taylor Brock-Fisher, Brittany A. Petros, Eric Y. Dai, Ariana T. Leonelli, Sabrina T. Dobbins, Katelyn S. Messer, Audrey B. Nathanson, Amelia Capone, Nancy Littlehale, Viola Appiah-Danquah, Siang Dim, Gage K. Moreno, Maura Crowther, Kannon A. Lee, Katherine C. DeRuff, Bronwyn L. MacInnis, Michael Springer, Pardis C. Sabeti, Kathryn E. Stephenson

## Abstract

**Background:** Households are a major setting for SARS-CoV-2 infections, but there remains a lack of knowledge regarding the dynamics of viral transmission, particularly in the setting of widespread pre-existing SARS-CoV-2 immunity and evolving variants.

**Methods:** We conducted a prospective, case-ascertained household transmission study in the greater Boston area in March-July 2022. Anterior nasal swabs, along with clinical and demographic data, were collected for 14 days. Nasal swabs were tested for SARS-CoV-2 by PCR. Whole genome sequencing was performed on high-titer samples.

**Results:** We enrolled 33 households in a primary analysis set, with a median age of participants of 25 years old (range 2-66); 98% of whom had received at least 2 doses of a COVID-19 vaccine. 58% of households had a secondary case during follow up and the secondary attack rate (SAR) for contacts infected was 39%. We further examined a strict analysis set of 21 households that had only 1 PCR+ case at baseline, finding an SAR of 22.5%. Genomic epidemiology further determined that there were multiple sources of infection for household contacts, including the index case and outside introductions. When limiting estimates to only highly probable transmissions given epidemiologic and genomic data, the SAR was 18.4%.

**Conclusions:** Household contacts of a person newly diagnosed with COVID-19 are at high risk for SARS-CoV-2 infection in the following 2 weeks. This is, however, not only due to infection from the household index case, but also because the presence of an infected household member implies increased SARS-CoV-2 community transmission. Further studies to understand and mitigate household transmission are needed.

**Key Points:** When community transmission of SARS-CoV-2 is high, distinguishing household transmissions from independent introductions is difficult with either epidemiologic or genomic data alone. Here, we conducted daily nasal sampling with genomic sequencing to understand the dynamics of viral transmission within households.

## INTRODUCTION

In March 2022, the SARS-CoV-2 Omicron variant BA.2 and its descendants caused multiple outbreaks across the greater Boston area of Massachusetts^1^. More than 95% of the population of Massachusetts at that time had received at least one dose of a COVID-19 vaccine, and a large number of individuals had been infected with the Omicron BA.1 variant only a few months prior^1^. Despite this presence of partial population immunity, there were over 200,000 new COVID-19 cases reported in Massachusetts between March and July 2022, coincident with the dropping of mask requirements and a reduction in social distancing policies.

Throughout the COVID-19 pandemic, households have been a major setting for SARS-CoV-2 transmission^2^. A meta-analysis of 135 studies published in 2022 found that the secondary attack rate (SAR) – the risk of transmission from index case to household contact – of exposed household contacts was 30-36% during the Alpha and Delta SARS-CoV-2 waves and 43% during the early (BA.1) Omicron period^2^. Factors associated with transmission risk were the vaccine status of the exposed contact, household density (i.e., the number of people sleeping in the same room), and immunological factors such as the magnitude of cross-reactive memory T cells to SARS-CoV-2 antigens^3-5^. In general, the SARs reported in households, and other residential settings^6^, have been far higher than reported in other settings such as schools, where a SAR of 2.8% was reported for exposed students in 10 Massachusetts schools during the Delta wave^7^.

Despite the large number and range of household studies performed to date, there remains a lack of precision and detail regarding the dynamics of SARS-CoV-2 household transmission. These studies are each individually small in size, and most lack careful characterization on the genomic level. This uncertainty is particularly important in the increasingly common scenario where there is (1) widespread pre-existing SARS-CoV-2 immunity, either from vaccination or infection, (2) emergence of new SARS-CoV-2 strains that are partially immune evasive, and (3) few infection control precautions in home and community environments. Knowing the risk of household transmission in this scenario, as well as identifying factors that can predict lower SARs, would enhance our ability to counsel patients appropriately and meaningfully reduce SARS-CoV-2 infections in households.

To address this research gap, we conducted a household SARS-CoV-2 transmission study in the greater Boston area in March-July 2022 in which we used clinical, demographic, viral kinetic, and genomic data for high-resolution analyses of SARS-CoV-2 transmission dynamics.

## METHODS

We conducted a prospective, observational, case-ascertained transmission study of household contacts of index SARS-CoV-2 cases identified in the greater Boston and Chelsea areas of Massachusetts in the United States. The primary recruitment pathway was via ambulatory COVID-19 testing sites associated with Beth Israel Deaconess Medical Center (BIDMC).

### Ethics

The BIDMC Committee on Clinical Investigation, its Institutional Review Board (IRB), approved the study protocol (#2022P000021). Consent procedures were completed via phone/video conferencing or in-person. All enrolled participants provided verbal informed consent. Participants who met applicable IRB guidelines for provision of assent underwent an age-appropriate assent process; informed consent for study participation was obtained from each participant’s parent or legal guardian before any study-specific procedures were performed. Sequencing of SARS-CoV-2-positive specimens was covered under protocol #1612793224, reviewed and approved by the Massachusetts Institute of Technology (MIT) IRB.

### Study Population

An index case was eligible for inclusion if the participant (1) had a positive SARS-CoV-2 test either by rapid diagnostic assay or RT-PCR no more than 5 days prior to enrollment and had no more than 5 days of acute respiratory illness symptoms prior to testing, and (2) lived, and had plans to live in his/her household for the follow-up period of 14 days, and (3) was not hospitalized and had not been hospitalized since the date of illness onset, and (4) was at least 2 years old. An index case was excluded from the study if the participant did not live in a household (e.g., lived alone or in a congregate setting), or if they reported that any other person in the household had an acute respiratory illness or tested positive for SARS-CoV-2 in the 7 days before the test date or illness onset date of the index case.

A household contact was eligible for inclusion if the participant (1) routinely slept (at least half of the nights in the last month) in the same household as the index case, and (2) slept in the household at least once between 1 day prior to the earliest of the index case’s illness onset or positive test date and (3) had plans to live in the household for the follow-up period, and (4) was at least 2 years old.

### Respiratory Specimen Collection

Each enrolled index case and household member self-collected an anterior nasal swab (Rhinostics dry swab in a sterile tube; https://rhinostics.com/) on each day of the 14-day follow-up period. The swab was self-collected regardless of whether the individual had acute respiratory illness signs or symptoms. Samples were dropped off by the participants at the BIDMC Clinical Research Center or the Chelsea clinic. A subset of participants who were treated with nirmatrelvir-ritonavir were asked to extend the follow-up period and continue self-swabbing for an additional 7 days in order to monitor for potential virologic rebound^8^.

### Questionnaires

At enrollment, each consenting household member and index case, or his/her parent/guardian, was asked to provide information on participant demographics, recent exposures to COVID-19, presence of high-risk conditions, prior and current symptoms of COVID-19, COVID-19 vaccination status, household characteristics, interactions with other enrolled and non-enrolled household members and people outside the home, and other social or medical history, including other vaccination history, as deemed appropriate. An optional follow-up questionnaire was administered to participants on days 7 and 14.

### SARS-CoV-2 Viral Load Quantification

Viral loads were measured with the Quaeris SARS-CoV-2 Assay, a real-time reverse transcription polymerase chain reaction (rRT–PCR) test, using the Luna Probe One-Step RT-qPCR Kit (No ROX) [NEB E3007]^9^. The SARS-CoV-2 primer and probe set detects RNA from the SARS-CoV-2 N1 and RdRP genes, and uses the human RNase P gene as a positive control. When received by the laboratory, samples were rehydrated with 300 µl phosphate buffered saline (PBS), inactivated at 65°C, and subsequently used directly as input for the Quaeris assay without extraction. rRT-PCR was performed on an Applied Biosystem QuantStudio 7 instrument (software version 1.7). Liquid handling was automated using either the Tecan Fluent 1080, the Hamilton Star, or the Multidrop combi dispenser. N1 gene cycle threshold (Ct) values are reported. Viral rebound was defined as at least 2 negative (Ct≥35) PCR results followed by at least 2 positive (Ct<35) results.

### SARS-CoV-2 Whole Genome Sequencing

Anterior nasal swabs that were positive for SARS-CoV-2 with Ct values of maximally 32 via the Quaris assay were sequenced as previously described^10^. Briefly, following inactivation with Buffer AVL, RNA was extracted from the samples using the MagMAX mirVana Total RNA Isolation Kit for the Kingfisher Flex (Thermo Fisher #A27828) according to the manufacturer’s instructions. The ARTIC v4.1 primer set was used to amplify SARS-CoV-2 genetic material, from which Illumina DNA Prep sequencing libraries were prepared and sequenced on the NextSeq 550^11^.

### SARS-CoV-2 Genomic Data Analysis

Sequencing reads were demultiplexed, filtered to remove adapter and contaminant sequences, depleted of reads mapping to the human genome, and assembled by alignment to the reference sequence NC045512.2 via the viral-ngs v2.1.33 pipeline^12^. The following optional inputs were used with the assemble_refbased workflow to accommodate amplicon-based sequencing: major_cutoff = 0.5, min_coverage = 20, skip_mark_dupes = TRUE, and trim_coords_bed = “gs://pathogen-public-dbs/v1/amplicon_primers-ARTICv4.1_NC_045512.2.bed”.

Lineages were assigned to viral genomes using Pango v4.1.3 pango-data v1.17^13^. Consensus single nucleotide variants (SNVs) were determined after removing ambiguous sites and sites prone to amplicon sequencing error^14^. Intrahost single nucleotide variants (iSNVs) were called using LoFreq and were filtered as follows: (i) masking of known problematic sites in SARS-CoV-2 genome, (ii) site read depth of minimally 100; (iii) allele frequency of minimally 3%; and (iv) no evidence of strand bias (via Fisher’s exact test with p > 0.05)^11,15^.

### Transmission Analysis

To infer the relatedness of cases within a household, the most complete genome (i.e., the genome with the highest percentage of unambiguous sites) was used for each person with a minimum unambiguous genome length of 15,000 bp. The genetic (SNV) distance between pairs of cases was used alongside the serial interval, viral kinetics, iSNV information, and test status of other individuals at baseline to classify households into transmission categories. All pairs of cases within each household were assessed, excluding those inconsistent with the longitudinal testing data.

Transmissions were categorized as highly probable, possible, or unlikely. Pairs with a genetic distance of less than 2 SNVs and an iSNV in a putative donor that reached consensus in the putative recipient (i.e., transmission of a minor variant through a tight transmission bottleneck) were considered highly probable^16,17^. An index-contact pair in the strict analysis set (i.e., from a household with a single infected participant and documented PCR-contacts at baseline) with a genetic distance of less than 3 SNVs and a serial interval of less than 7 days were also considered highly probable transmissions. Pairs with a genetic distance of less than 3 SNVs and a serial interval of less than 7 days, but without iSNV support, were categorized as a possible transmission, though we cannot rule out a non-household recent common ancestor (i.e.., a shared exposure)^18,19^. Pairs with a genetic distance of 3 or more SNVs or a serial interval of minimally 7 days were considered unlikely. Highly probable and possible transmission links were plotted.^20,21^

### Phylogenetic Analysis

Contextual genomes (10 BA.1 sequences each from Massachusetts and the United States, and 50 BA.2 sequences each from Massachusetts and the United States, isolated between 2022-02-01 and 2022-08-01) were downloaded from NCBI GenBank. These sequences, along with the most complete genome assembled per study participant (of minimally 15,000 bp), were aligned to the reference sequence (NC_045512.2) using Nextclade v.2.14.1^22^. A phylogenetic tree was estimated using IQ-TREE v.2.2.2.6^23^ and was visualized and annotated using ggtree v.3.8.2^24^ in R v.4.3.1.

## RESULTS

### Screening, Enrollment, and Follow-up

Enrollment began on March 3, 2022 and continued until July 9, 2022, after local SARS-CoV-2 case counts had declined^1^. We enrolled 38 households containing 38 index cases and 77 household contacts; Figure 1. One household was subsequently found to be ineligible and 4 households did not complete 14 days of follow up. The remaining 33 households (33 index cases and 66 household contacts) constituted our primary analysis set; 85% of these households (28/33) had all household members participate. Of the 33 households in the primary analysis set, 3 households had an index case that was PCR-at the time of first research swab, and 9 households had at least one household contact that were PCR+ at the time of first research swab. The remaining 21 households (21 index cases and 40 PCR-household contacts) constituted our strict analysis set. The median number of persons per household in the primary analysis set was 3.5 (range, 2-5; Supplementary Table 2). The median house size was 1400 square feet (range, 740-3300; Supplementary Table 2).

**Figure 1.**
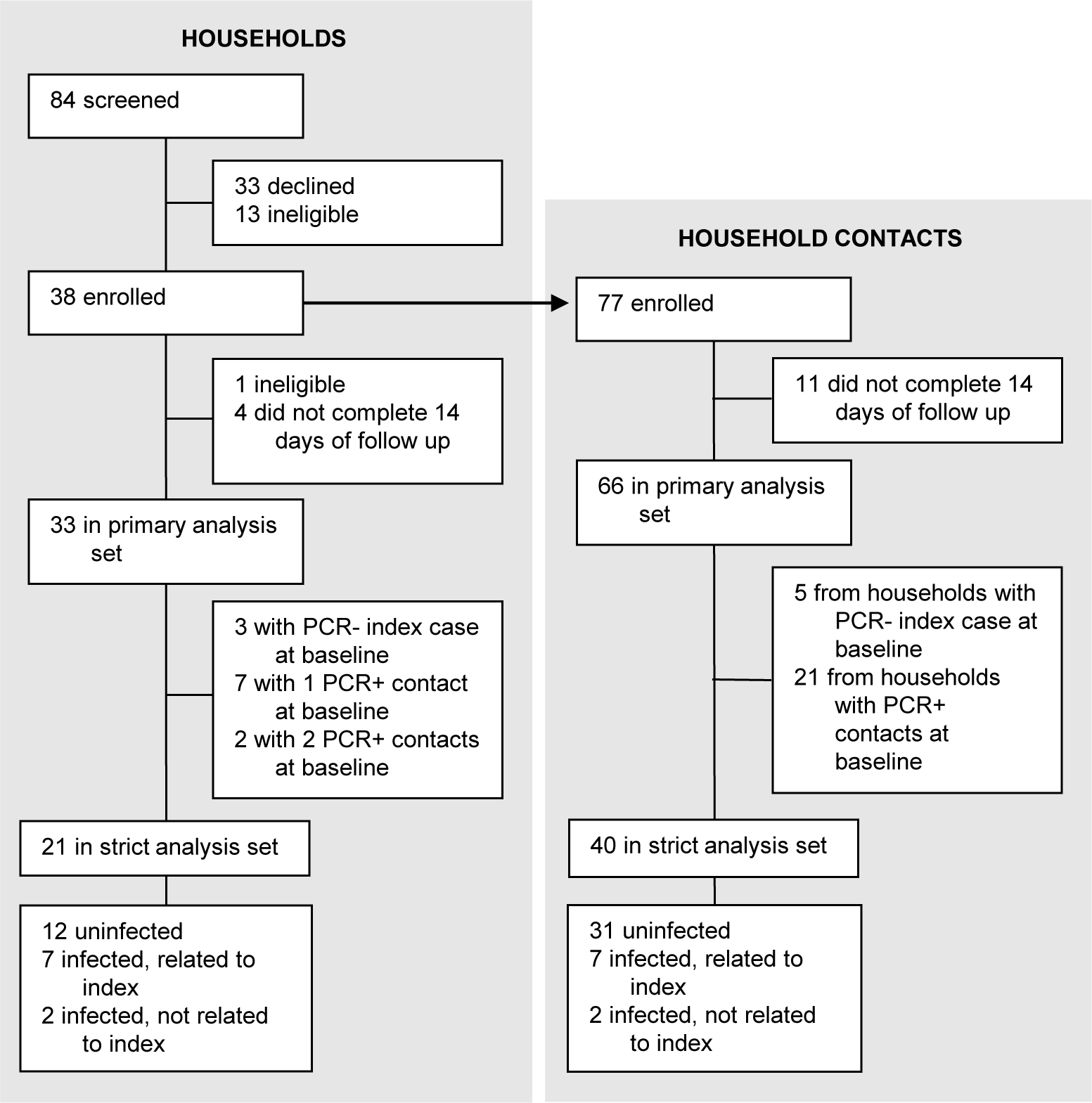
Trial profile.

### Study Population

Demographic and clinical characteristics of index cases and household contacts in the primary analysis set are shown in Table 1 and Supplemental Table 1. The median age was 25 years (range, 5-64) for index cases and 39 years (range, 2-66) for household contacts. 23% of participants were children in elementary or middle school. Participants were predominantly White and Non-Hispanic. 81% of participants reported having no medical conditions. The most common reported medical conditions were asthma (12%), autoimmune or immune system disease (8%), and high blood pressure (4%). 10% of participants reported having COVID-19 at some point prior to the study (though symptom- and testing-based ascertainment bias is likely). 98% of participants received at least 2 vaccine doses. The median duration between most recent vaccination or most recent infection was 160 days (range 0-307).

**Table 1.** Baseline Characteristics of Study Population.

| **Characteristic** | **All Participants**<br>**(N=99)** | **Index Cases**<br>**(N=33)** | **Contacts**<br>**(N=66)** |
| --- | --- | --- | --- |
| **Age** (Median, Min-Max) | 27 (2-66) | 25 (5-64) | 39 (2-66) |
| **Sex assigned at birth** (N, %) – |  |  |  |
| Male | 44 (48.4) | 14 (43.8) | 30 (50.9) |
| Female | 47 (51.7) | 18 (56.3) | 29 (49.2) |
| Missing | 8 | 1 | 7 |
| **Race** (N, %) – |  |  |  |
| American Indian or Alaskan Native | 10 (11.0) | 3 (9.4) | 7 (11.9) |
| Asian | 2 (2.2) | 1 (3.1) | 1 (1.7) |
| Black or African American | 0 | 0 | 0 |
| Native Hawaiian or Other Pacific Islander | 0 | 0 | 0 |
| White | 79 (86.8) | 28 (87.5) | 51 (86.4) |
| Multiple | 0 | 0 | 0 |
| Prefer Not to Report | 0 | 0 | 0 |
| Other | 0 | 0 | 0 |
| Missing | 8 | 1 | 7 |
| **Ethnicity** (N, %) – |  |  |  |
| Hispanic or Latino / Latinx | 3 (3.3) | 1 (3.1) | 2 (3.4) |
| Not Hispanic or Latino / Latinx | 88 (96.7) | 31 (96.9) | 57 (96.6) |
| Missing | 8 | 1 | 7 |
| **Body Mass Index** (Median, Min/Max) | 22.9 (14.1-7.2) | 23.0 (16.1-37.1) | 22.7 (14.1-47.2) |
| Missing | 19 | 9 | 10 |
| **Medical Conditions** (N, %) |  |  |  |
| None | 69 (81.2) | 21 (72.4) | 48 (85.7) |
| Cancer | 0 | 0 | 0 |
| Diabetes | 1 (1.2) | 1 (3.5) | 0 |
| Asthma or wheezing | 10 (11.8) | 5 (17.2) | 5 (8.9) |
| COPD/Emphysema | 0 | 0 | 0 |
| High blood pressure | 3 (3.5) | 1 (3.5) | 2 (3.6) |
| Coronary artery disease | 0 | 0 | 0 |
| Congestive heart failure | 0 | 0 | 0 |
| Peripheral artery disease | 0 | 0 | 0 |
| Other* | 2 (2.4) | 1 (3.5) | 1 (1.8) |
| Missing | 14 | 4 | 10 |
| **Autoimmune or Immune System Diseases, Any\*\*** (N, %) | 7 (8.2) | 1 (3.7) | 6 (10.3) |
| Missing | 14 | 6 | 8 |
| **Prior Diagnosis of COVID-19** (N, %) | 9 (9.5) | 1 (3.2) | 8 (12.5) |
| Missing | 4 | 2 | 2 |
| Days between most recent diagnosis and enrollment (Median, Min-Max) | 59 (18-780) | 780 | 57 (18-462) |
| Hospitalized for COVID-19 (N, %) – *missing 1* | 0 | 0 | 0 |
| **Prior Receipt of a COVID-19 Vaccine (N, %) – *missing 2*** | 95 (97.9) | 32 (100) | 63 (96.9) |
| Number of doses (Median, Min-Max) | 3 (2-4) | 3 (2-4) | 3 (2-4) |
| Days between most recent COVID-19 vaccination and enrollment (Median, Min-Max) – *missing 4* | 165 (1-308) | 165 (48-308) | 165 (1-307) |
| Days between most recent diagnosis OR most recent COVID-19 vaccination (whichever is more recent) and enrollment (Median, Min-Max) | 160 (0-308) | 164.5 (0-308) | 160 (1-307) |
| Missing | 2 | 1 | 1 |

\*Reported by participant: “heart aneurysm” (1); history of stroke (2)
\*\*Reported by participant: “irritable bowel syndrome, eczema and allergies” (1); “vitiligo” (1); “diverticulosis” (1); “Reynaud’s disease” (1)

### SARS-CoV-2 Infections

Of the 33 households in the primary analysis set, 19 (58%) had a household contact who tested positive by PCR during follow up (Figure 2A and Table 2). Of the 66 household contacts in the primary analysis set, 26 tested positive, indicating a putative SAR of 39%; 11 of these cases were detected on day 1 of swabbing. If limiting analysis to the 21 households in the strict analysis set, 9 (43%) households had a new incident infection during follow up, corresponding to 9 infections out of 40 household contacts, or a putative SAR of 22.5%.

**Figure 2.**
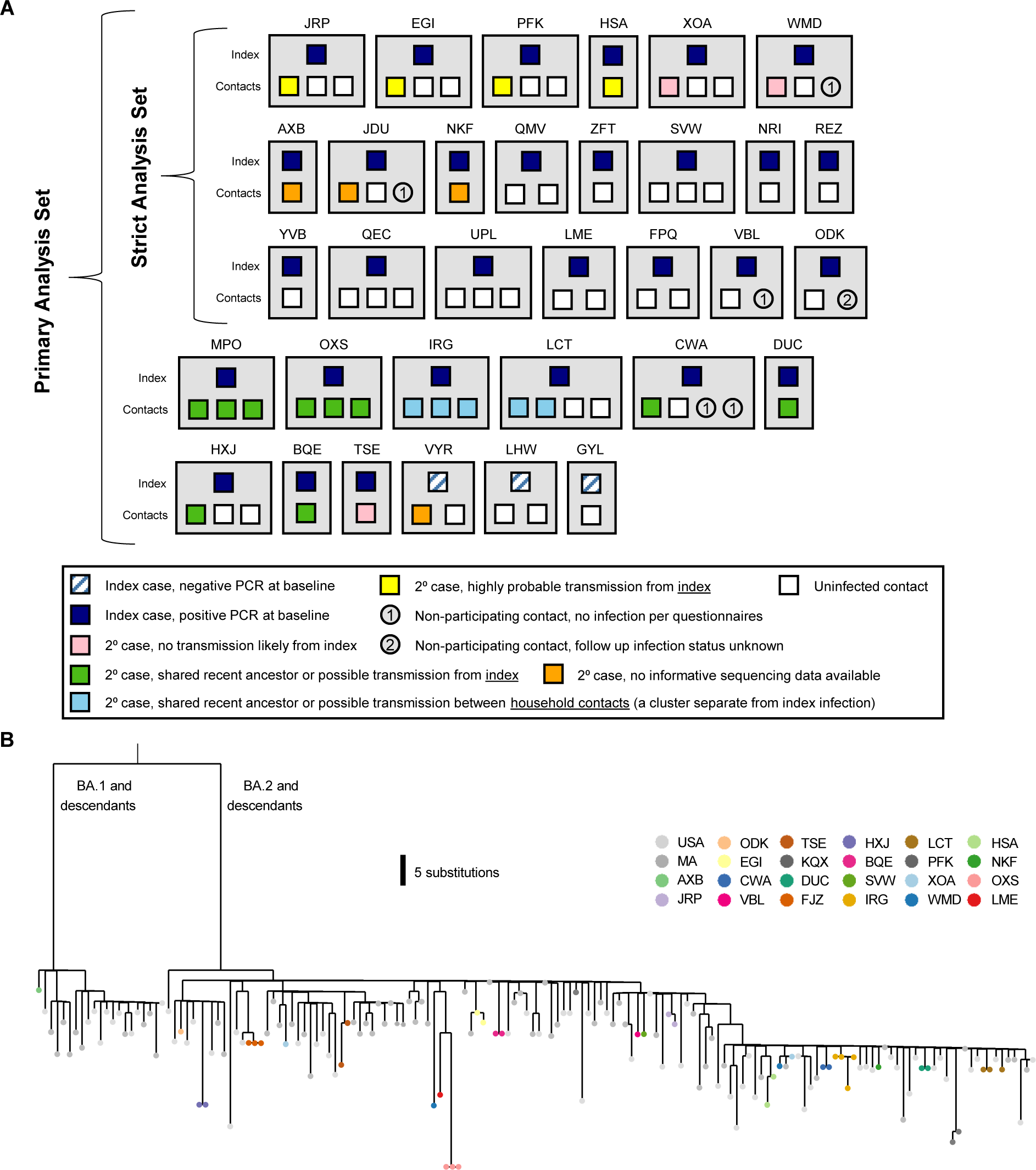
Household diagram. (A) Individual households are labeled with a 3-letter identifier and represented with grey boxes. Households in both the primary and strict analysis sets are shown. Each member of the household is shown within the grey box, with their outcome color-coded as described in the legend. (B) Phylogenetic map of positive samples.

**Table 2.** SARS-CoV-2 Secondary Attack Rates.

|  | **# Events** | **Total Households** | **Total Contacts** | **SAR (%)** |
| --- | --- | --- | --- | --- |
| **Primary Analysis Set** |  |  |  |  |
| Households with 2<sup>o</sup> cases | 19 | 33 | - | 57.6 |
| Individual 2<sup>o</sup> cases | 26 | - | 66 | 39.4 |
| **Strict Analysis Set** |  |  |  |  |
| Households with 2<sup>o</sup> cases | 9 | 21 | - | 42.9 |
| Households with 2<sup>o</sup> cases, excluding households with outside infection | 7 | 19 | - | 36.8 |
| Individual 2<sup>o</sup> cases | 9 | - | 40 | 22.5 |
| Individual 2<sup>o</sup> cases, excluding individuals with outside infection | 7 | - | 38 | 18.4 |

In the primary analysis set of 33 households, the median minimal Ct - corresponding to peak viral load - was 25.65 for index cases and 23 for infected household contacts (Supplemental Table 3). Median duration of viral shedding was 6.5 days and 8 days for index cases and contacts, respectively, and the median time to positive SARS-CoV-2 test from index diagnosis date was 3 days. The lower viral load and decreased duration of shedding for index cases is most likely due to the fact that cases were detected partway through infection. Eleven participants reported taking nirmatrelvir-ritonavir; 3 participants had viral rebound (Supplemental Figure 1A: MPO-02, BQE-01 and BQE-02)^8^.

### SARS-CoV-2 Sequencing

For the primary analysis set, approximately 1300 total anterior nasal swab samples were collected and analyzed via the rRT-PCR Quaris assay. 181 of these samples, from 48 unique individuals across 26 unique households, had a diagnostic Ct under 32 (mean = 30.1) and were thus subject to genomic sequencing. 178/181 (98.3%) samples had a nonzero unambiguous assembly length with a median length of 29,321bp. 143 samples, which came from 44 unique individuals among 24 unique households, produced partial genomes of at least 15,000 bp in length (Supplemental Table 4). 15 households yielded partial genomes from multiple individuals within the household. The vast majority of the sequences were BA.2 descendants, consistent with national trends (Figure 2B), though a BA.1 sequence was collected early in enrollment.

Focusing first on analyzable households in the strict analysis set (N=6), we found that four households had viral sequences that were either identical between index and secondary cases or differed by only 1-2 SNVs (Figure 3A, Supplemental Table 5). In these cases, transmission from index to secondary case was considered highly probable. In two households, we identified significant viral variation between sequences isolated from the index and secondary contacts, strongly suggesting that the secondary cases were independent introductions (Figure 3B). In household XOA, the index case was infected with a BA.2.13 variant, while the contact was infected with BA.2.12.1; there was a 13 SNV difference between sequences. In household WMD, the index case was infected with BA.2.12.1, and the contact was infected with BA.2; there was a 20 SNV difference between sequences. (Of the 3 remaining households with secondary cases in the strict analysis set (AXB, JDU and NKF), sequences were not available or couldn’t be analyzed; viral kinetic curves for these households are in Figure 3C). When excluding the 2 household contacts that were infected outside of the household, the SAR among individual household contacts in the strict analysis set was 18.4% (Table 2).

**Figure 3.**
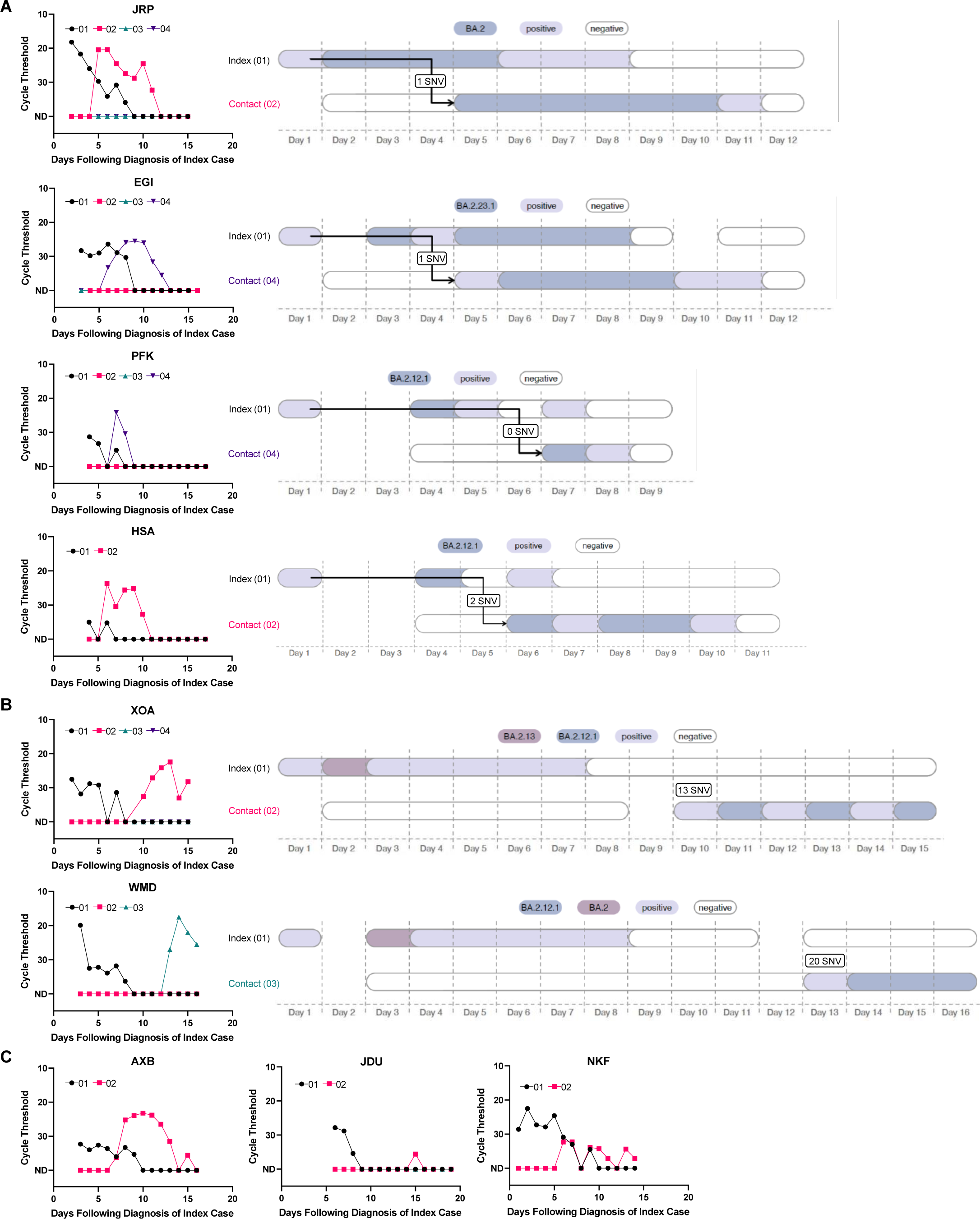
Viral load curves and transmission plots by household in the strict analysis set. (A) Households where transmission was determined to be highly probable are shown. (B) Households where transmission was determined to be unlikely are shown. (C) Households that did not yield sufficient sequencing data are shown.

Analyzing transmission networks among all the households in the primary analysis set revealed several interesting patterns. In 5 households, there were clusters of infections (2-3 per household) that were all genetically linked to the index case (Figure 4A (a representative example), and Supplemental Figure 1A). In these households, there were 0 SNVs different between index and contact sequences, but the close overlap in diagnosis dates suggests that either a shared common exposure or intra-household transmission was possible. In 3 households, there were two distinct clusters of infections detected (Figure 4B (a representative example), and Supplemental Figure 1B). In these households, the index case was infected with a virus that was 3-4 SNVs different than the viruses that circulated in the rest of the household. In 1 household (Figure 4C), there was evidence that the index case was co-infected with 2 contemporaneously circulating strains (BA.2.1.12 and BA.2.10) while the contact was infected with solely BA.2.1.12; given the close diagnosis dates, this could be explained by either a shared social network or transmission with a bottleneck.

**Figure 4.**
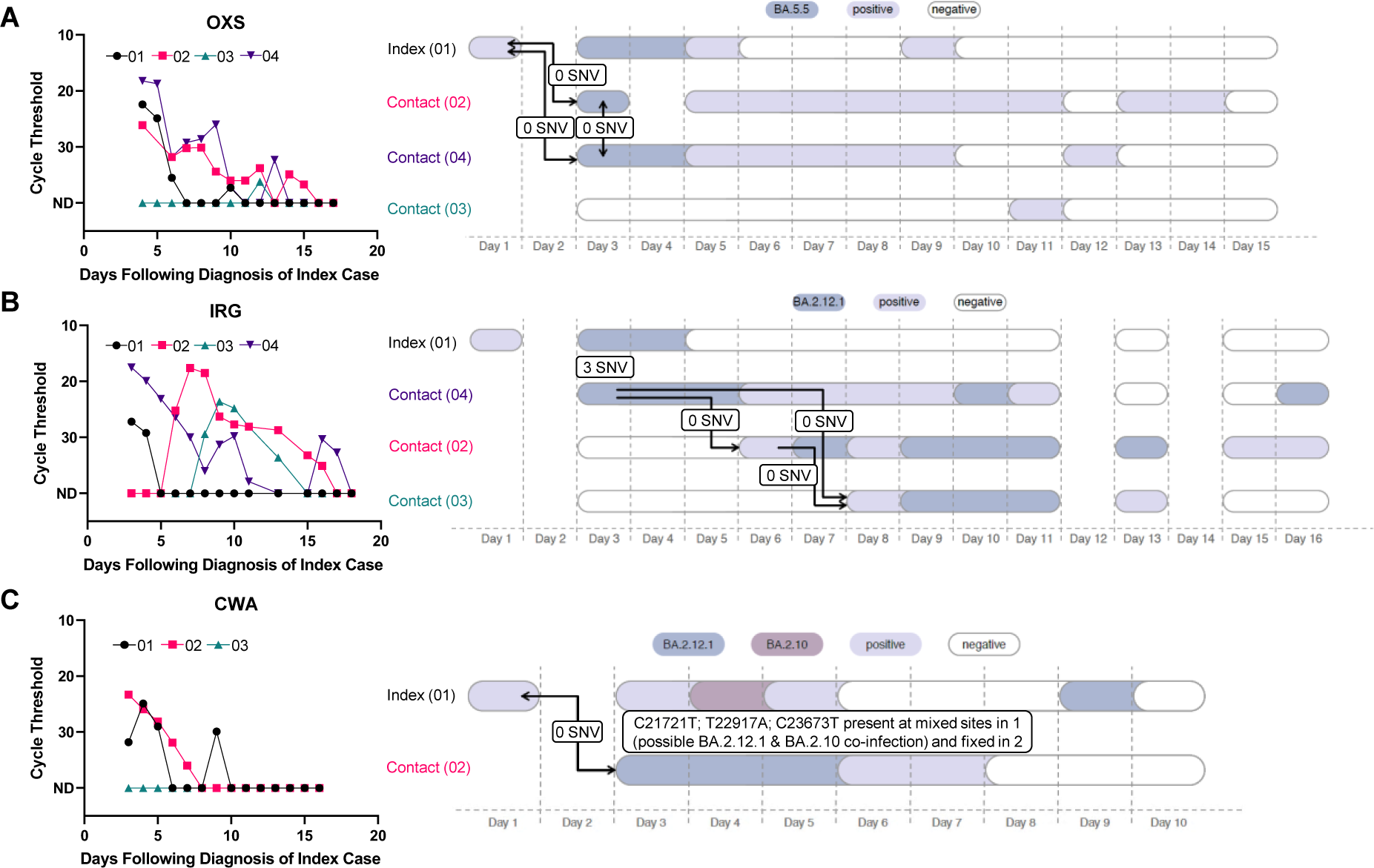
Representative viral load curves and transmission plots from primary analysis set. (A) A representative household is shown where either a shared exposure (i.e., social network) exists or within-household transmission has occurred, as all cases are genetically linked to the index case. (B) A representative household is shown where two separate infection clusters were identified. (C) A household is shown where a putative co-infection was identified in the index case.

### Potential Predictors of Transmission

To explore potential predictors of transmission between an index case and household contact, we examined the 7 household contacts where transmission from the index was highly likely compared to the 28 participating contacts that remained uninfected. Using Wilcoxon rank sum and Fisher’s exact tests (Table 3), we found that there was no association between infection status and the contact’s prior COVID-19 vaccination, number of prior vaccines received, history of prior infection, days since vaccination and/or infection, age or sex assigned at birth. There was also no association between infection status and the associated index case’s prior COVID-19 vaccine status, minimum Ct value, or duration of SARS-CoV-2 shedding, and no association between infection status and the number of persons in the household or and square footage. The small sample size and homogeneity of many of these variables (e.g., vaccine history) limited the power of our analysis to detect predictors of transmission risk.

**Table 3.** Household Contact Characteristics by Infection Status (Strict Analysis Set), Excluding 2^0^ Cases with Outside Infections.

|  | **Infected Contact**<br>**(N=7)** | **Not Infected Contact**<br>**(N=28)** | **p-value** |
| --- | --- | --- | --- |
| **Characteristic of Household Contact** |  |  |  |
| Age (Median, Min-Max) | 46 (13-50) | 28.5 (10-64) | 0.76<sup>a</sup> |
| Male sex assigned at birth (N, %)<br>Missing | 2 (33.3)<br>1 | 13 (59.1%)<br>6 | 0.37<sup>b</sup> |
| Number of COVID-19 vaccines received (Median, Min-Max)<br>Missing | 3 (2-3)<br>0 | 3 (2-4)<br>1 | 1.00<sup>b</sup> |
| Days from last infection/vaccination (Median, Min-Max)<br>Missing | 146 (119-193)<br>0 | 138 (-1-197)<br>6 | 0.45<sup>a</sup> |
| **Characteristic of Associated Index Case** |  |  |  |
| Lowest Ct value (Median, Min-Max) | 27.8 (18.2-35) | 25 (15.9-33.9) | 0.19<sup>a</sup> |
| Duration of shedding in days (Median, Min-Max)<br>Missing | 7 (3-9)<br>0 | 8 (1-15)<br>0 | 0.41<sup>a</sup> |
| **Characteristic of Associated Household** |  |  |  |
| Number of persons in household (Median, Min-Max)<br>Missing | 4 (2-4)<br>0 | 4 (2-4)<br>1 | 0.10<sup>b</sup> |
| Square footage (Median, Min-Max)<br>Missing | 1140 (740-3300)<br>2 | 1000 (740-3300)<br>15 | 0.59<sup>a</sup> |
| a. Wilcoxon rank sum test<br>b. Fisher's Exact |  |  |  |

## DISCUSSION

Here we studied the transmission dynamics of SARS-CoV-2 within households using prospective daily PCR surveillance, clinical and demographic data, and genomic epidemiology. Our study is well-positioned to reflect the transmission dynamics of households consisting of relatively young, healthy individuals with pre-existing immunity to SARS-CoV-2. It was conducted during a wave of Omicron BA.2 and its sub-lineages. Our study population consisted primarily of families with 2-5 members; 23% of our participants were children in elementary or middle school. Participants were highly vaccinated, having received a median of 3 COVID-19 vaccines. Only 10% had a known prior COVID-19 diagnosis and the population was overall very healthy.

Our study found that the risk of SARS-CoV-2 in exposed households is very high, consistent with previous reports. In our primary analysis set, we found that 58% of enrolled households went on to have a second SARS-CoV-2 case in the household over the next 2 weeks. Nearly a third of households had a positive secondary case identified on the very first day of testing. At the individual level, we found that 39% of household contacts tested positive for SARS-CoV-2 during follow up. Following the infection of a household contact, putatively negative household members must make decisions around travel and attending school or work. Here, we provide data that could inform this value-based decision.

We further aimed to more rigorously estimate the risk of SARS-CoV-2 transmission directly from an index case, performing a strict analysis including only households that had one confirmed PCR+ index case and only PCR-household contacts at the beginning of surveillance. In this analysis, we identified 9 cases among 40 household contacts, or a SAR of 22.5%. Using genomic epidemiology, we then excluded 2 cases as these were likely acquired outside of the household. We thus determined that SAR was 18.4%. Our study was conducted when multiple BA.2 descendants were circulating, and this genetic diversity improved our ability to rule out putative transmission events.

Using genomic epidemiology across all the households, we found that the infectious source of secondary cases was varied. The most common source of infection was the index case, but other sources included an imported case (outside the household), shared exposures with the index, or other infected contacts within the home. Our findings, particularly evidence of shared exposures and imported cases, highlight that infection within a household serves as a proxy for broader risk for community transmission, emphasizing the importance of maintaining vigilant public health measures both inside and outside of the home. More public health attention should be focused on mitigating this risk via vaccination and therapeutic advances, enhanced and early case detection and isolation, and non-pharmaceutical interventions such as masking and ventilation.

SAR estimates have varied considerably across multiple studies, due to varying study designs, populations, and circulating SARS-CoV-2 variants. During the Alpha and Delta waves, the SAR for exposed household contacts was reported between 30-36%, rising to 43% during the early (BA.1) Omicron period^2^. Here we report that during a BA.2 wave (and its descendants), the SAR was 39% using a larger primary set of households, 22.5% with a strict set of households, and 18.4% when further incorporating genomic data. This lower estimate using genomic data is likely a more accurate reflection of the true risk of household exposure.

Our small sample size substantially limited our power to assess predictors of transmission, especially because prior COVID-19 vaccination was near universal in our study cohort. Given the complexity of estimating the magnitude of pre-existing SARS-CoV-2 immunity by history alone, particularly in the context of both infection and prior vaccines, future household transmission studies would benefit from larger cohorts and baseline immunologic assessments of humoral, cellular and mucosal immune responses.

In summary, we determined that the risk of household transmission of SARS-CoV-2 is very high, even when using genomic epidemiology to exclude imported cases. Further interventions to block household transmission should be studied, especially in the now-common scenario when population immunity and vaccine coverage are high but viral variants are increasingly immune evasive.

## Data Availability

All data produced in the present study are available upon reasonable request to the authors

## Acknowledgements

We thank the participants and staff at the Center for Virology and Vaccine Research Clinical Trials Unit, the Harvard Catalyst Clinical Research Center, and the Beth Israel Deaconess Primary Care—Chelsea Clinic. We also thank Daniel J. Park for providing important feedback on the manuscript, and Janet Morgan for administrative support.

---

**Supplemental Figure 1.**
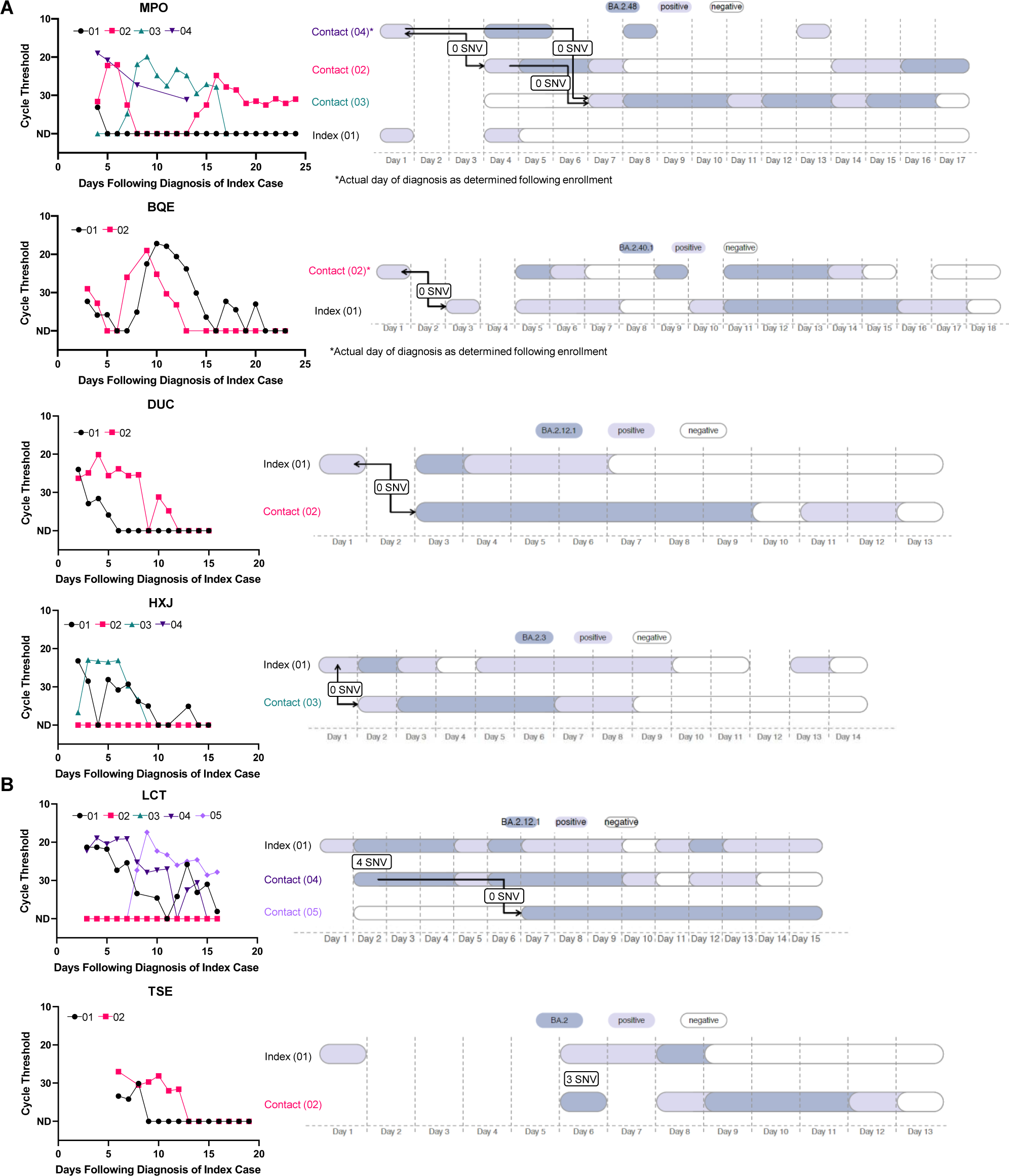
Additional viral load curves and transmission plots by household in the primary analysis set. Households are shown where either a shared exposure exists or possible transmission has occurred, as all cases are genetically linked to the index case. Households are shown where two separate infection clusters were identified.

## SUPPLEMENTAL TABLES

**Supplemental Table 1.** Additional Baseline Data.

| **Characteristic** | **All Participants**<br>**(N=99)** | **Index Cases**<br>**(N=33)** | **Contacts**<br>**(N=66)** |
| --- | --- | --- | --- |
| **Currently Working / Employment (N, %) – *missing 13*** |  |  |  |
| Yes | 57 (66.3) | 18 (69.2) | 39 (65.0) |
| No | 29 (33.7) | 8 (30.8) | 21 (35.0) |
| Missing | 13 | 7 | 6 |
| **Student (N, %) – *missing 9*** |  |  |  |
| Yes | 32 (35.6) | 13 (43.3) | 19 (31.7) |
| No | 58 (64.4) | 17 (56.7) | 41 (68.3) |
| Missing | 9 | 3 | 6 |
| **If yes (N=32), level (N, %)** |  |  |  |
| Elementary | 11 (34.4) | 5 (38.5) | 6 (31.6) |
| Middle School | 12 (37.5) | 5 (38.5) | 7 (36.8) |
| High School | 5 (15.6) | 2 (15.4) | 3 (15.8) |
| Community College | 0 | 0 | 0 |
| College or University | 4 (12.5) | 1 (7.7) | 3 (15.8) |
| **Currently Uses eCigarettes/Vape (N, %) – *missing 9*** | 1 (1.1) | 0 | 1 (1.6) |
| **Currently Smokes Cigarettes (N, %) – *missing 9*** | 1 (1.1) | 0 | 1 (1.6) |
| **History of Smoking Cigarettes (N, %) – *missing 10*** | 11 (12.4) | 3 (10.3) | 8 (13.3) |
| If yes, packs per day (N, %) |  |  |  |
| 0.5 | 7 (63.6) | 1 (33.3) | 6 (75.0) |
| 1.0 | 4 (36.4) | 2 (66.7) | 2 (25.0) |
| 1.5 | 0 | 0 | 0 |
| 2.0 | 0 | 0 | 0 |
| 2.5 | 0 | 0 | 0 |
| 3 or more | 0 | 0 | 0 |
| If yes, how many years (N, %) |  |  |  |
| 1-5 | 8 (72.7) | 2 (66.7) | 6 (75.0) |
| 5-10 | 0 | 0 | 0 |
| 10-15 | 1 (9.1) | 0 | 1 (12.5) |
| 15-20 | 0 | 0 | 0 |
| 20 or more | 2 (18.2) | 1 (33.3) | 1 (12.5) |
| **Concurrent Use of Steroids or Immune Suppressing Medications (N, %) – *missing 12*** | 6 (6.9) | 1 (3.6) | 5 (8.5) |
| **Concurrent use of NSAIDs – *missing 13*** | 38 (44.2) | 10 (37.0) | 28 (47.5) |

**Supplemental Table 2.**
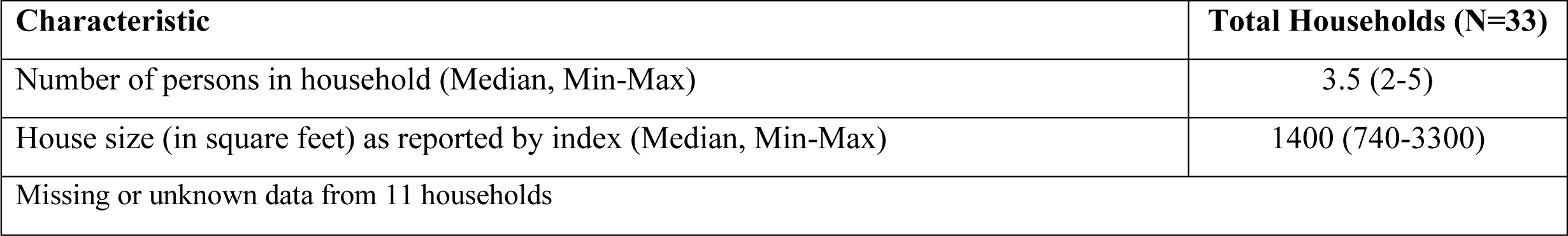
Baseline Characteristics of Study Households in Primary Analysis Set.

**Supplemental Table 3.** SARS-CoV-2 Viral Kinetics in Primary Analysis Set, Excluding 2 Cases of Outside Introduction.

| **Outcome** | **Index Cases**<br>**(N=30)<sup>a</sup>** | **Infected Contacts**<br>**(N=26)** |
| --- | --- | --- |
| Lowest Ct (median, min-max) | 25.65 (15.9-35) | 23 (8.1-36.2) |
| Duration of viral shedding<sup>b</sup> (median, min-max) | 6.5 days (1-19) | 8 days (1-20) |
| Time to first positive SARS-CoV-2 from index diagnosis date | N/A | 3 days (0-10) |

a. Excluding 3 index cases who were PCR negative at baseline.
b. From first positive SARS-CoV-2 test.

**Supplemental Table 4.** Sequencing Performance.

| **Household** | **Participant** | **Samples** | **Samples Producing Genome with Minimum Unambiguous Genome Length of 15 kbp** |
| --- | --- | --- | --- |
| AXB | 1 | 1 | 0 |
|  | 2 | 6 | 5 |
| JRP | 1 | 5 | 5 |
|  | 2 | 6 | 6 |
| ODK | 1 | 1 | 1 |
| EGI | 1 | 6 | 6 |
|  | 4 | 5 | 4 |
| CWA | 1 | 4 | 3 |
|  | 2 | 4 | 3 |
| VBL | 1 | 1 | 1 |
| TSE | 1 | 1 | 1 |
|  | 2 | 6 | 5 |
| KQX | 1 | 1 | 1 |
| DUC | 1 | 2 | 1 |
|  | 2 | 8 | 7 |
| MPO | 2 | 5 | 4 |
|  | 3 | 9 | 7 |
|  | 4 | 4 | 3 |
| HXJ | 1 | 4 | 1 |
|  | 3 | 5 | 5 |
| BQE | 1 | 7 | 5 |
|  | 2 | 5 | 5 |
| SVW | 1 | 1 | 1 |
| IRG | 1 | 2 | 2 |
|  | 2 | 6 | 6 |
|  | 3 | 4 | 3 |
|  | 4 | 8 | 5 |
| LCT | 1 | 7 | 5 |
|  | 4 | 11 | 9 |
|  | 5 | 9 | 9 |
| PFK | 1 | 1 | 1 |
|  | 4 | 2 | 1 |
| JDU | 1 | 1 | 1 |
| XOA | 1 | 6 | 1 |
|  | 2 | 4 | 4 |
| REZ | 1 | 1 | 0 |
| WMD | 1 | 2 | 1 |
|  | 3 | 4 | 3 |
| HSA | 1 | 1 | 1 |
|  | 2 | 5 | 3 |
| YVB | 1 | 1 | 0 |
| NKF | 1 | 1 | 1 |
|  | 2 | 1 | 0 |
| QEC | 1 | 1 | 1 |
| OXS | 1 | 2 | 2 |
|  | 2 | 1 | 1 |
|  | 4 | 2 | 2 |
| LME | 1 | 1 | 1 |

**Supplemental Table 5.**
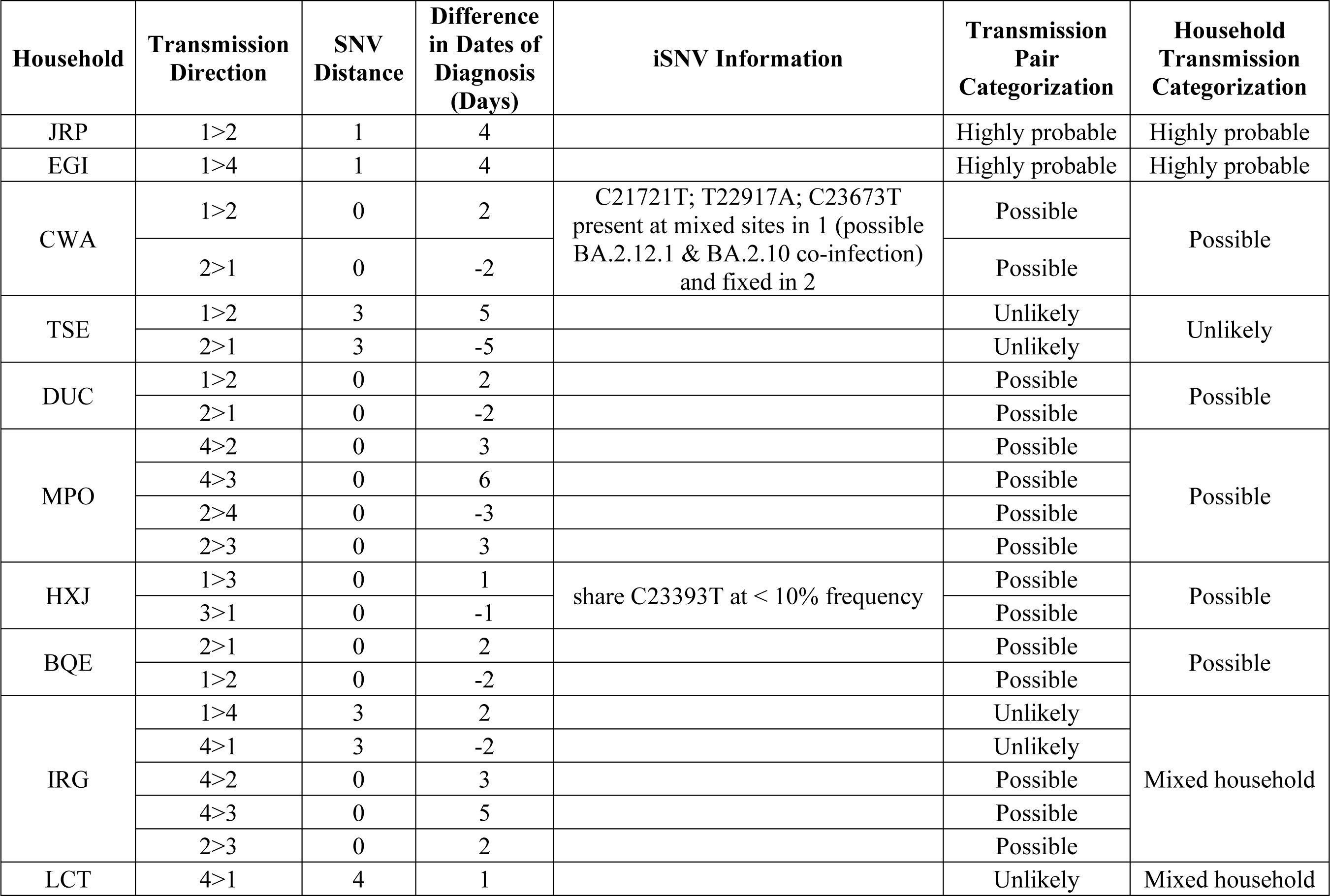
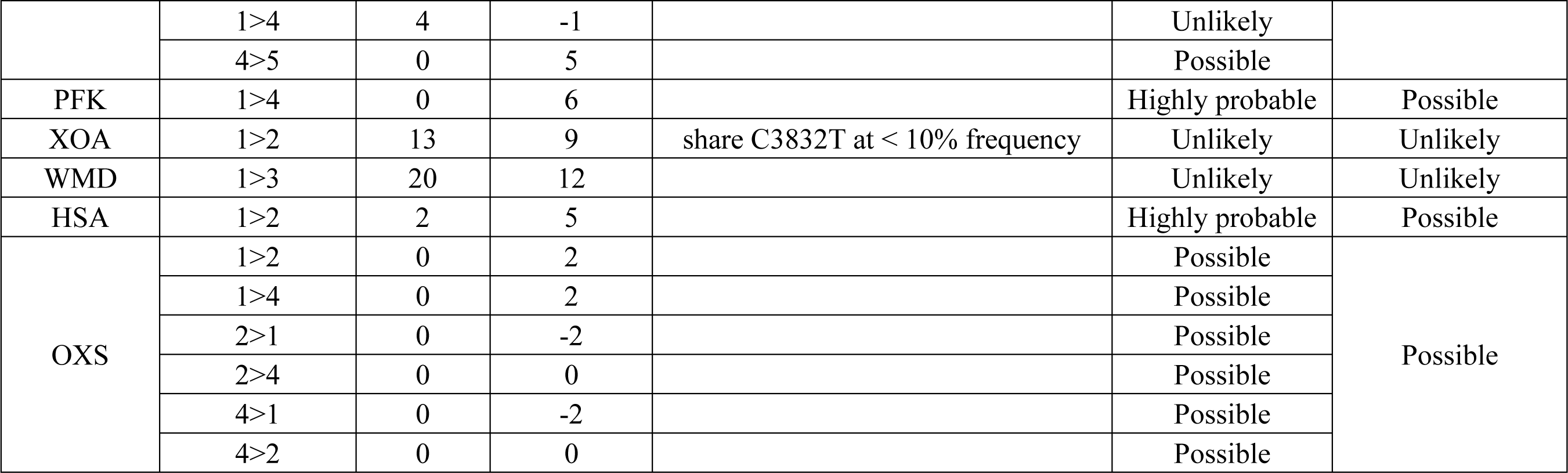
Household Transmission Analysis.

## References

1. Massachusetts Department of Public Health COVID-19 Dashboard, www.mass.gov.

2. Madewell ZJ, Yang Y, Longini IM, Jr., Halloran ME, Dean NE. Household Secondary Attack Rates of SARS-CoV-2 by Variant and Vaccination Status: An Updated Systematic Review and Meta-analysis. JAMA Netw Open. Apr 1 2022;5(4):e229317. doi:10.1001/jamanetworkopen.2022.9317

3. Singanayagam A, Hakki S, Dunning J, et al. Community transmission and viral load kinetics of the SARS-CoV-2 delta (B.1.617.2) variant in vaccinated and unvaccinated individuals in the UK: a prospective, longitudinal, cohort study. Lancet Infect Dis. Feb 2022;22(2):183-195. doi:10.1016/S1473-3099(21)00648-4

4. Kleynhans J, Walaza S, Martinson NA, et al. Household Transmission of Severe Acute Respiratory Syndrome Coronavirus 2 From Adult Index Cases With and Without Human Immunodeficiency Virus in South Africa, 2020-2021: A Case-Ascertained, Prospective, Observational Household Transmission Study. Clin Infect Dis. Feb 8 2023;76(3):e71-e81. doi:10.1093/cid/ciac640

5. Kundu R, Narean JS, Wang L, et al. Cross-reactive memory T cells associate with protection against SARS-CoV-2 infection in COVID-19 contacts. Nat Commun. Jan 10 2022;13(1):80. doi:10.1038/s41467-021-27674-x

6. Petros BA, Turcinovic J, Welch NL, et al. Early Introduction and Rise of the Omicron Severe Acute Respiratory Syndrome Coronavirus 2 (SARS-CoV-2) Variant in Highly Vaccinated University Populations. Clin Infect Dis. Feb 8 2023;76(3):e400–e408. doi:10.1093/cid/ciac413

7. Nelson SB, Dugdale CM, Brenner IR, et al. Prevalence and Risk Factors for School-Associated Transmission of SARS-CoV-2. JAMA Health Forum. Aug 4 2023;4(8):e232310. doi:10.1001/jamahealthforum.2023.2310

8. Dai EY LK, Nathanson AB, Leonelli AT, Petros BA, Brock-Fisher T, Dobbins ST, MacInnis BL, Capone A, Littlehale N, Boucau J, Marino C, Barczak AK, Sabeti PC, Springer M, Stephenson KE. Viral Kinetics of Severe Acute Respiratory Syndrome Coronavirus 2 (SARS-CoV-2) Omicron Infection in mRNA-Vaccinated Individuals Treated and Not Treated with Nirmatrelvir-Ritonavir. medRxiv [Preprint]. medRxiv 2022011322269257. 2022;

9. Emergency Use Authorization (EUA) Summary Quaeris SARS-CoV-2 Assay. Available at: https://wwwfdagov/media/149445/download. 2021;

10. Petros BA, Paull JS, Tomkins-Tinch CH, et al. Multimodal surveillance of SARS-CoV-2 at a university enables development of a robust outbreak response framework. Med. Dec 9 2022;3(12):883–900 e13. doi:10.1016/j.medj.2022.09.003

11. https://github.com/artic-network/artic-ncov2019/tree/master/primer_schemes/nCoV-2019/V4.1.

12. Park, D et al., https://github.com/broadinstitute/viral-ngs/tree/v1.25.0.

13. O’Toole A, Scher E, Underwood A, et al. Assignment of epidemiological lineages in an emerging pandemic using the pangolin tool. Virus Evol. 2021;7(2):veab064. doi:10.1093/ve/veab064

14. https://github.com/W-L/ProblematicSites_SARS-CoV2/blob/master/problematic_sites_sarsCov2.vcf.

15. Wilm A, Aw PP, Bertrand D, et al. LoFreq: a sequence-quality aware, ultra-sensitive variant caller for uncovering cell-population heterogeneity from high-throughput sequencing datasets. Nucleic Acids Res. Dec 2012;40(22):11189-201. doi:10.1093/nar/gks918

16. Braun KM, Moreno GK, Wagner C, et al. Acute SARS-CoV-2 infections harbor limited within-host diversity and transmit via tight transmission bottlenecks. PLoS Pathog. Aug 2021;17(8):e1009849. doi:10.1371/journal.ppat.1009849

17. Song JS, Lee J, Kim M, et al. Serial Intervals and Household Transmission of SARS-CoV-2 Omicron Variant, South Korea, 2021. Emerg Infect Dis. Mar 2022;28(3):756-759. doi:10.3201/eid2803.212607

18. Bendall EE, Callear AP, Getz A, et al. Rapid transmission and tight bottlenecks constrain the evolution of highly transmissible SARS-CoV-2 variants. Nat Commun. Jan 17 2023;14(1):272. doi:10.1038/s41467-023-36001-5

19. Lythgoe KA, Hall M, Ferretti L, et al. SARS-CoV-2 within-host diversity and transmission. Science. Apr 16 2021;372(6539)doi:10.1126/science.abg0821

20. Campbell F, Cori A, Ferguson N, Jombart T. Bayesian inference of transmission chains using timing of symptoms, pathogen genomes and contact data. PLoS Comput Biol. Mar 2019;15(3):e1006930. doi:10.1371/journal.pcbi.1006930

21. Jombart T, Cori A, Didelot X, Cauchemez S, Fraser C, Ferguson N. Bayesian reconstruction of disease outbreaks by combining epidemiologic and genomic data. PLoS Comput Biol. Jan 2014;10(1):e1003457. doi:10.1371/journal.pcbi.1003457

22. Aksamentov I, Roemer C, Hodcroft EB, Neher RA. Nextclade: clade assignment, mutation calling and quality control for viral genomes. The Journal of Open Source Software. 2021;6(67)

23. Nguyen LT, Schmidt HA, von Haeseler A, Minh BQ. IQ-TREE: a fast and effective stochastic algorithm for estimating maximum-likelihood phylogenies. Mol Biol Evol. Jan 2015;32(1):268–74. doi:10.1093/molbev/msu300

24. Yu G, Smith DK, Zhu H, Guan Y, Tsan-Yuk Lam T. ggtree: an r package for visualization and annotation of phylogenetic trees with their covariates and other associated data. Methods in Ecology and Evolution. 2016;

